# Endovascular Thrombectomy for Acute Stroke with Large Ischemic Core: A Meta-analysis of Randomized Clinical Trials

**DOI:** 10.1101/2024.10.07.24314988

**Authors:** Chun-Hsien Lin, Meng Lee, Bruce Ovbiagele, David S Liebeskind, Borja Sanz-Cuesta, Jeffrey L Saver

## Abstract

**Background:** Endovascular thrombectomy (EVT) is standard treatment for an acute ischemic stroke due to large-vessel occlusions (AIS-LVO) with a small ischemic core, but the effect of EVT on AIS-LVO with a large ischemic core remains unclear. The objective was to assess the benefit and safety of EVT for AIS-LVO with large ischemic cores.

**Methods and Finding:** PubMed, the Cochrane Central Register of Controlled Trials, and ClinicalTrials.gov were searched from January 1 2000 to September 25 2025. Randomized clinical trials (RCTs) of patients with AIS-LVO and large ischemic cores that compared EVT plus medical care vs. medical care alone were evaluated. Risk ratio (RR) with 95% CI was used to measure outcomes of EVT vs. medical care alone. Primary outcome was functional independence, defined as modified Rankin Scale (mRS) of 0 to 2 at 90 days; and lead secondary outcome was reduced disability, defined as ordinal shift of mRS. Safety outcomes were requiring constant care or death (mRS 5 to 6), death, and early symptomatic intracranial hemorrhage (sICH). We included 6 RCTs comprising 1870 patients (women 826 [44.2%]) with AIS-LVO. Pooled results showed that at 90 days, EVT vs. medical care alone was associated with greater functional independence (RR 2.53, 95% CI, 1.95-3.29; number needed to treat [NNT], 9) and reduced disability (common odds ratio 1.63, 95% CI 1.38-1.93; NNT, 4). EVT vs. medical care alone was associated with a lower risk of requring constant care or death (RR 0.74, 95% CI 0.66 to 0.84; NNT, 7). The rates of death and early sICH were not significantly different between the EVT and medical care alone groups.

**Conclusions:** EVT combined with medical care compared with medical care alone may be associated with improved functional outcomes and reduced severe disability or death at 90 days for acute stroke with large ischemic cores.

**PROSPERO registration number:** CRD42024514605

## INTRODUCTION

Endovascular thrombectomy (EVT) has been proven to be more effective than medical care alone in improving functional independence and reducing disability in patients with acute ischemic stroke due to large vessel occlusions (AIS-LVO) and small to moderate ischemic cores, as defined by an Alberta Stroke Program Early Computed Tomographic Score (ASPECTS) value of at least 6 or a an ischemic core volume less than 31 to 70 ml.^1–3^ However, about 20% of patients with AIS-LVO have large ischemic cores and these patients generally have very poor prognosis, including progression of stroke symptoms, requiring constant care, and death.^4^

Most randomized controlled trials enrolling patients with large ischemic cores indicated a benefit in functional outcome after EVT compared with medical care alone.^5–9^ However, one trial did not show significant benefits upon functional outcome.^10^ Meta-analysis of randomized clinical trials may help to resolve conflicting results of individual trials, increase precision of effect magnitude estimates, enhance generalizability, and inform clinical practice guidelines.^11^ We therefore conducted a systematic review of randomized clinical trials to explore the efficacy and safety of EVT compared with medical care alone in patients with AIS-LVO having large ischemic cores.

## METHODS

The Preferred Reporting Items for Systematic Reviews and Meta-analyses (PRISMA) reporting guideline was used for abstracting data and validity of this meta-analysis.^12^ The protocol was registered with PROSPERO(CRD42024514605).

### Search Methods and Resources

We searched PubMed, the Cochrane Central Register of Controlled Trials, and the clinical trial registry maintained at ClinicalTrials.gov for studies published from January 1, 2000, to September 25, 2024 using the following terms: stroke or cerebrovascular disease or brain ischemia or brain infarct or cerebrovascular accident and large core or large infarct or large ischemic and endovascular therapy or endovascular thrombectomy or endovascular treatment or mechanical thrombectomy or intra-arterial therapy. We restricted our search to randomized clinical trials. There were no language restrictions. We also reviewed the introduction and discussion sections of retrieved trials to identify additional trials. Two investigators (C.-H. L. and M.L.) independently screened and identified potential trials, and discrepancies were resolved by discussion with a third investigator (J.L.S).

### Study selection

Criteria for inclusion of a study were: (1) study design was a randomized clinical trial; (2) patients had AIS-LVO; (3) patients had large infarctions (e.g. ASPECT ≤ 5 or an estimated ischemic-core volume of 50 ml or greater on CT perfusion imaging); (4) trials compared EVT plus medical care vs medical care alone; and (5) functional independence, defined as modified Rankin Scale (mRS) score 0 to 2, at 90 days was reported as an endpoint. Studies were excluded if the study design was a registry, case report, case-control, or cohort. We extracted characteristics of each trial, which included patient age, sex, imaging criteria for enrollment, and outcomes assessed. Two investigators (C.-H. L. and M.L.) independently abstracted the data and any discrepant judgments were resolved by referencing the original report.

### Study Quality Assessment

Since all of the included studies were randomized controlled trials, we assessed the overall bias (e.g. bias arising from the randomization process, bias due to deviations from intended interventions, bias due to missing outcome data, bias in measurement of the outcome, and bias in selection of the reported result) by using the RoB-2 tool.^13^

### Outcomes

The primary outcome was functional independence (mRS score of 0 to 2) at 90 days. The secondary outcomes were reduced disability (ordinal shift across the range of mRS scores toward a better outcome, mRS 0-1/2/3/4/5/6) at 90 days, ambulation (mRS score of 0 to 3) at 90 days, being nondisabled (mRS score of 0 to 1) at 90 days, and early neurologic improvement. The safety outcomes were requiring constant care or death (defined as mRS score 5 to 6) at 90 days, death within 90 days, and early symptomatic intracranial hemorrhage. We specifically analyzed the outcome of mRS score or 5 to 6 because there is little difference along the disability dimension among mRS score of 5 and 6 outcomes^14^ and recent stroke expert consensus reports categorize mRS scores of 5 to 6 together as Very Poor outcome.^15^

### Statistical Analysis

The analysis plan was performed on an intention-to-treat basis. We computed the random-effects estimate based on the inverse variance method when 2 or more studies provided sufficient data for a given outcome. Risk ratio (RR) with 95% confidence interval (CI) was used as a measure of the association of EVT vs medical care alone with binary outcomes. Common odds ratio with 95% CI was used as a measure of the association of EVT vs medical care alone with an ordinal shift across the range of mRS scores toward a better outcome at 90 days. All reported P values were two-sided, with significance set at <0.05. Heterogeneity was assessed by a P value determined by the use of χ2 statistics and I^2^ statistics, and I^2^ values of 0% to 29%, 30% to 49%, 50% to 74%, and 75% to 100% consider to represent not important, moderate, substantial, and considerable inconsistency, respectively.^16^ Number needed to treat values were derived for dichotomous outcomes as the inverse of the absolute risk reduction and for mRS shift using the joint outcome table algorithmic min-max specification method.^17^

To identify any trial that might have exerted a disproportionate influence on the summary treatment effect, we conducted a sensitivity test by removing each individual trial from the meta-analysis one at a time.^11^ Publication bias was assessed visually by a funnel plot displaying standard error as the measure of sample size and risk ratio as the measure of treatment effect.^11^ Grading of Recommendations, Assessment, Development and Evaluations was used to evaluate summaries of evidence for the outcomes.^11,18^ The Cochrane Collaboration’s Review Manager Software Package (RevMan, version 5.4) was used for this meta-analysis.

## RESULTS

We identified 11 full articles for detailed assessment, of which 5 did not meet the inclusion criteria; therefore, the final analysis included 6 randomized clinical trials (eFigure 1 in Supplement).^5–10^ The characteristics of the included trials are shown in Table 1.^5–10^ Overall, 1870 patients (women 826 [44.2%]) with large ischemic strokes were enrolled with 940 been randomly assigned to EVT and 930 been randomly assigned to medical care alone. The Recovery by Endovascular Salvage for Cerebral Ultra-Acute Embolism–Japan Large Ischemic Core Trial (RESCUE-Japan LMIT),^5^ the Endovascular Therapy in Acute Anterior Circulation Large Vessel Occlusive Patients with a Large Infarct Core (ANGEL-ASPECT) trial,^6^ the Randomized Controlled Trial to Optimize Patient’s Selection for Endovascular Treatment in Acute Ischemic Stroke (SLELCT2),^9^ and the Thrombectomy for Emergent Salvage of Large Anterior Circulation Ischemic Stroke (TESLA) trial^10^ enrolled patients up to 24 hours, whereas the Efficacy and Safety of Thrombectomy in Stroke With Extended Lesion and Extended Time Window (TENSION) trial^7^ enrolled patients within 12 hours and the Large Stroke Therapy Evaluation (LASTE) trial^8^ enrolled patients within 6.5 hours of last known well. Mean age ranged from 66 years to 76 years.^5^ Median baseline NIHSS score ranged from 16^6^ to 22.^5^ Median ASPECTS value was 2 in the LASTE trial^8^ and 3 or 4 among other included trials. The TESLA trial^10^ used ASPECTS value of 2 to 5 on non-contrast CT to select patients while other trials also used advanced imaging modalities, such as MRI or perfusion imaging, to select patients. The RESCUE-Japan LIMIT^5^ used mRS value of 0 to 3 as the primary outcome, the ANGEL-ASPECT trial,^6^ the SELECT2,^9^ the TENSION trial,^7^ and the LASTE trial^8^ used an ordinal shift in the distribution of scores on the mRS as the primary outcome, and the TESLA trial^10^ used score on utility-weighted mRS as the primary outcome.

**Figure 1.**
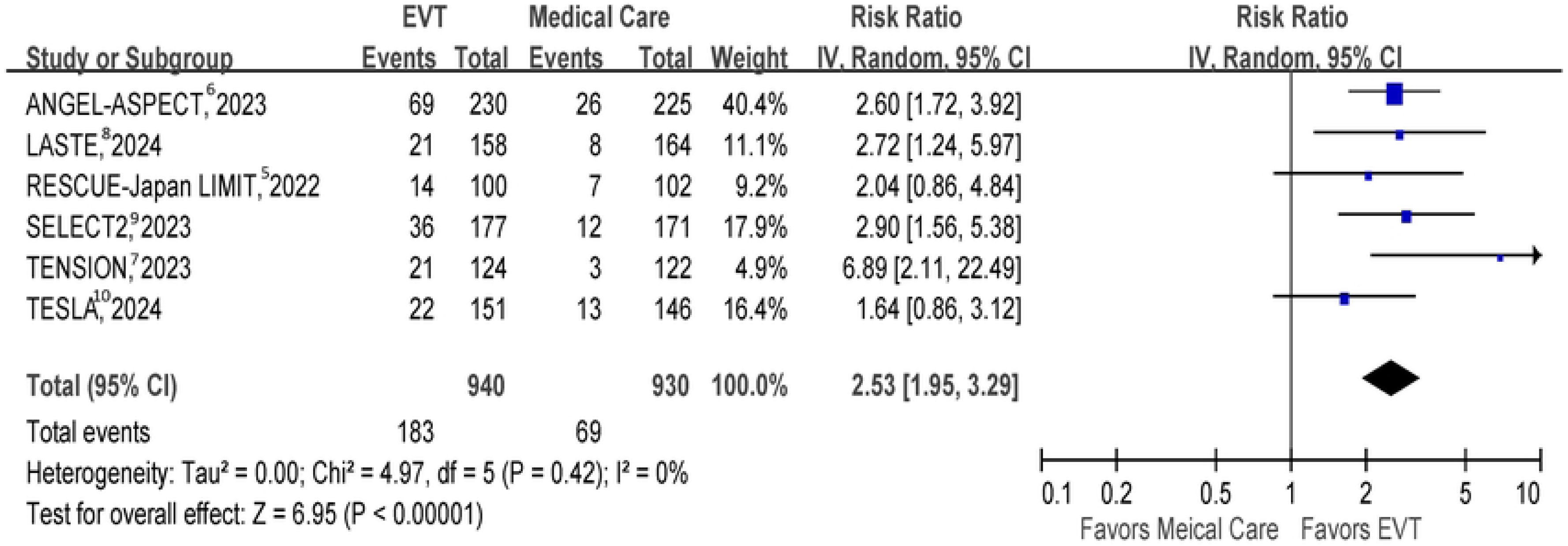
Functional Independence at 90 days. Risk ratio with 95% confidence interval of functional independence, defined as modified Rankin Scale 0 to 2, with EVT plus medical care compared with medical care alone in AIS-LVO patients with large ischemic cores at 90 days. AIS-LVO, acute ischemic stroke due to large-vessel occlusions; EVT, endovascular thrombectomy.

**Table 1.** Baseline characteristics of included trials.

|  | RESCUE-Japan<br>LIMIT,<sup>5</sup> 2022 | ANGEL-ASPECT,<sup>6</sup><br>2023 | SELECT2,<sup>9</sup> 2023 | TESLA,<sup>10</sup> 2024 | TENSION,<sup>7</sup> 2023 | LASTE,<sup>8</sup> 2024 |
| --- | --- | --- | --- | --- | --- | --- |
| Countries | Japan | China | USA, Canada,<br>Europe, Australia,<br>New Zealand | USA | Europe, Canada | France |
| Inclusion criteria | Age ≥ 18y, NIHSS ≥ 6, within 24h, occlusion of ICA or M1, ASPECT 3-5 on DWI or CT, pre-stroke mRS 0-1 | Age 18-80y, NIHSS 6-30, within 24h, occlusion of ICA or M1, ASPECT 3-5 on CT (no limit with infarct-core) or ASPECT 0-2 (infarct-core 70-100ml) or ASPECT >5 (infarct-core 70-100ml) between 6-24hr, pre-stroke mRS 0-1 | Age 18-85y, NIHSS ≥ 6, within 24h, occlusion of ICA or M1, ASPECT 3-5 on CT or core ≥ 50ml on CT perfusion, pre-stroke mRS 0-1 | Age 18-85y, NIHSS ≥ 6, within 24h, occlusion of ICA or M1, ASPECT 2-5 on NCCT, pre-stroke mRS 0-1 | Age ≥ 18y, NIHSS ≤ 26, within 12h, occlusion of ICA or M1, ASPECT 3-5 on DWI or CT, pre-stroke mRS 0-2 | Age > 18y, NIHSS ≥ 6, within 6.5h, occlusion of ICA or M1, ASPECT 0-5 on CT or MRI, pre-stroke mRS 0-1 |
| Sample size (% of women), EVT vs medical care | 101 (46) vs 102 (43) | 230 (41) vs 225 (36) | 178 (40) vs 174 (43) | 75 (49) vs 63 (43) | 125 (45) vs 128 (52) | 159 (48) vs 165 (47) |
| Age, y, EVT vs medical care | 77 vs 76 | 68 vs 67 | 66 vs 67 | 66 vs 68 | 73 vs 74 | 73 vs 74 |
| Median NIHSS, EVT vs medical care | 22 vs 22 | 16 vs 15 | 19 vs 19 | 19 vs 18 | 19 vs 18 | 21 vs 21 |
| **Occlusion site, n (%), EVT vs medical care** |  |  |  |  |  |  |
| ICA | 47 (47) vs 49 (48) | 83 (36) vs 81 (36) | 80 (45) vs 66 (38) | 37 (24) vs 31 (21) | 41 (33) vs 37 (29) | 69 (43) vs 74 (45) |
| M1 segment | 74 (73) vs 70 (69) | 145 (63) vs 142 (63) | 91 (51) vs 100 (58) | 119 (78) vs 124 (84) | 83 (66) vs 88(69) | 88 (55) vs 91 (55) |
| ASPECTS value, median, EVT vs medical care | 3 vs 4 | 3 vs 3 | 4 vs 4 | 4 vs 4 | 4 vs 4 | 2 vs 2 |
| Infarct volume, ml, median, EVT vs medical care | 94 vs 110 | 61 vs 63 | 82 vs 79 | NA | NA | 132 vs 137 |
| IVT, n (%), EVT vs medical care | 27 (27) vs 29 (28) | 66 (29) vs 63 (28) | 37 (21) vs 30 (17) | 31 (20) vs 30 (20) | 49 (39) vs 44 (34) | 55 (35) vs 58 (35) |
| Interval between onset and randomization, min, median, EVT vs medical care | 229 vs 214 | 453 vs 463 | 587 vs 544 | 652 vs 754 | 120 vs 126 | 271 vs 268 |

Acronym of trials: RESCUE Japan LIMIT, Recovery by Endovascular Salvage for Cerebral Ultra-Acute Embolism–Japan Large Ischemic Core Trial; ANGEL ASPECT, Endovascular Therapy in Acute Anterior Circulation Large Vessel Occlusive Patients with a Large Infarct Core; SELECT2, Randomized Controlled Trial to Optimize Patient's Selection for Endovascular Treatment in Acute Ischemic Stroke; TESLA, Thrombectomy for Emergent Salvage of Large Anterior Circulation Ischemic Stroke; TENSION, Efficacy and Safety of Thrombectomy in Stroke With Extended Lesion and Extended Time Window; LASTE, Large Stroke Therapy Evaluation.
AF, atrial fibrillation; ASPECTS, Alberta Stroke Program Early Computed Tomography Score; CT, Computed Tomography; DWI, diffuse weighted imaging; EVT, endovascular thrombectomy; ICA, internal carotid artery; IVT, intravenous thrombolysis; M1, M1 segment of middle cerebral artery; mRS, modified Rankin Scale; NA, not available; NCCT, noncontrast computed tomography; NIHSS, National Institutes of Health Stroke Scale; sICH, symptomatic intracranial hemorrhage; TICI, Thrombolysis in Cerebral Infarction

**Table 2.**
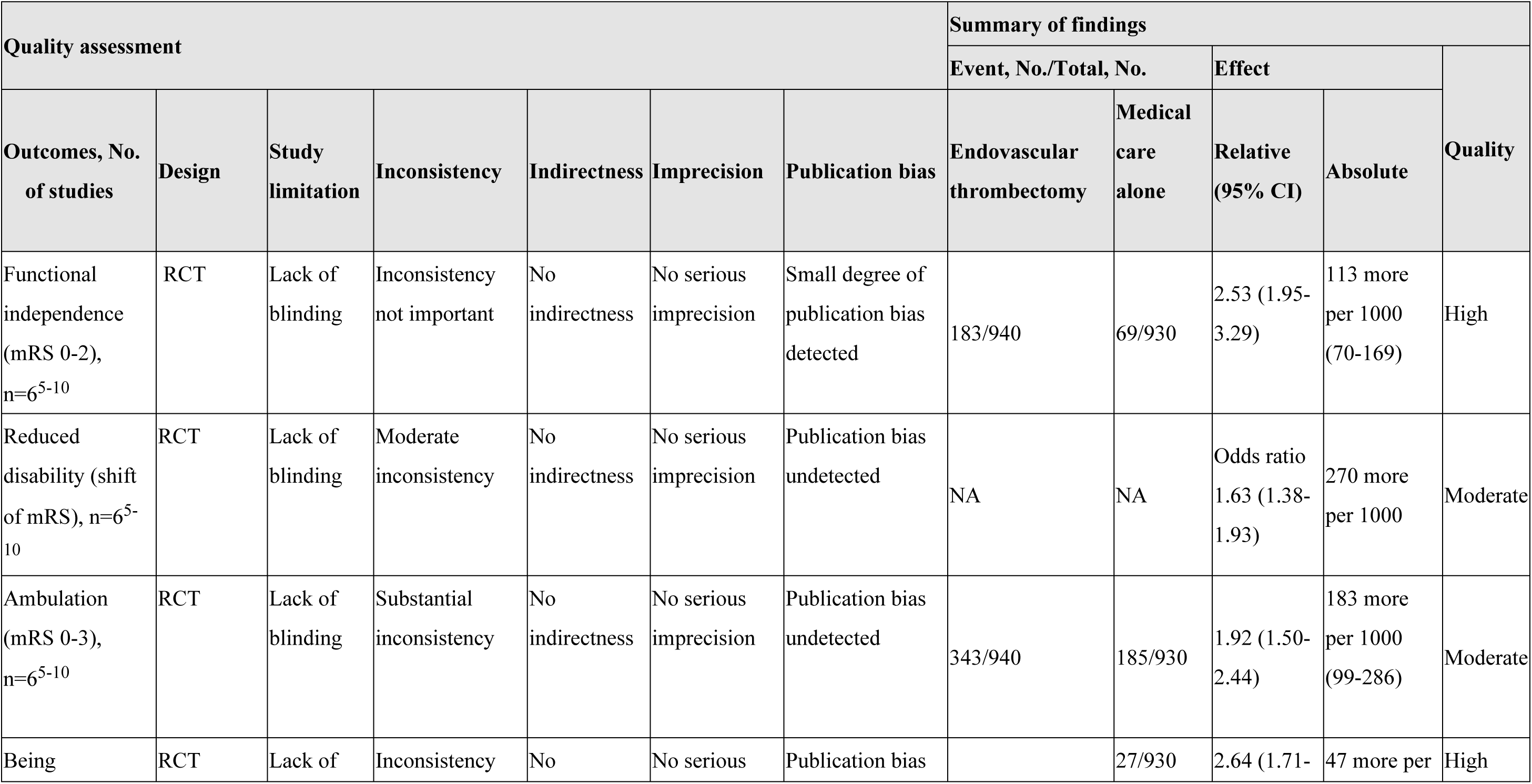
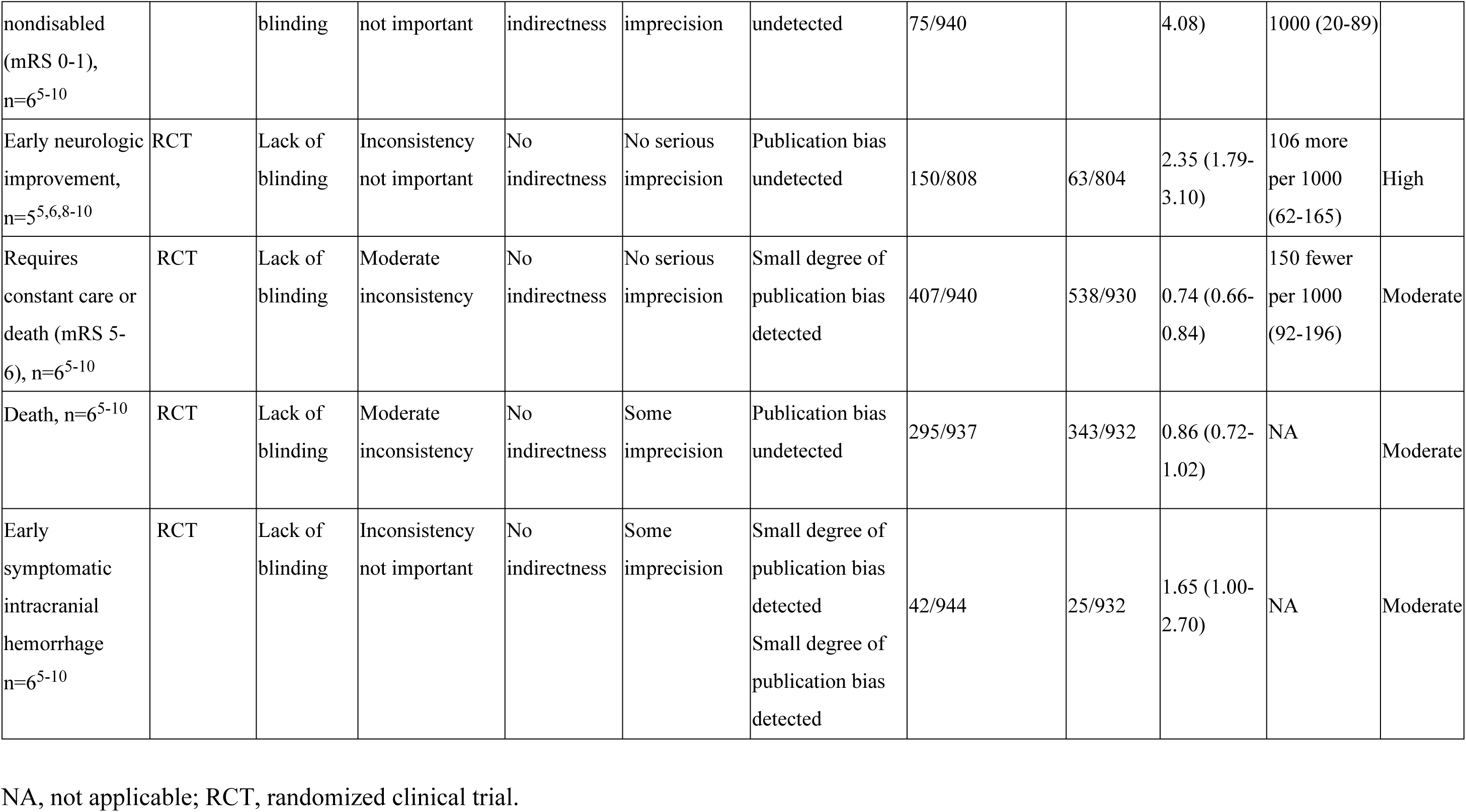
Grading of Recommendations, Assessment, Development and Evaluations (GRADE) for Outcomes.

Successful reperfusion, defined as Thrombolysis in Cerebral Infarction (TICI) score of 2b or 3, was achieved in 73% of cases in the TESLA trial’s EVT group.^10^ This rate was lower compared to other EVT trials, which reported successful reperfusion rates of 80% or higher.^5–8^ The RoB-2 for the included trials is summarized in eTable in Supplement.

### Primary Outcome

#### Functional Independence (mRS score of 0 to 2) at 90 days

Pooled results from the random-effects model showed that EVT compared with medical care alone was associated with a higher proportion of patients with functional independence at 90 days in AIS-LVO patients with large ischemic cores (6 trials; absolute risk, 19.5% vs 7.4%; RR 2.53, 95% CI 1.95 to 3.29, P < .001; number needed to treat [NNT], 9).^5–10^ This represented 113 more independent patients per 1000 treated with EVT compared with medical care alone. Heterogeneity was not important among included trials (*P* for heterogeneity = .42; *I*^2^ = 0%) (Figure 1).

### Secondary Outcomes

#### Reduced disability (ordinal shift of mRS) at 90 days

The distribution of mRS scores in the EVT and medical care groups is shown in Figure 2. Across these trials, allocation to EVT compared with medical care was associated with a higher proportion of patients achieving favorable outcomes at 90 days in AIS-LVO patients with large ischemic cores, including a directionally favorable effect at each of the 5 health state transitions over the mRS 0-1/2/3/4/5/6 range (Figure 2). Pooled results showed that EVT compared with medical care alone was associated with reduced disability at 90 days (6 trials; common odds ratio 1.63, 95% CI 1.38 to 1.93, P < .001; NNT, 4).^5–10^ This represented 270 more independent patients per 1000 treated with EVT compared with medical care alone. There was moderate heterogeneity among included trials (*P* for heterogeneity = .11; *I*^2^ = 45%) (eFigure 2 in Supplement).

**Figure 2.**
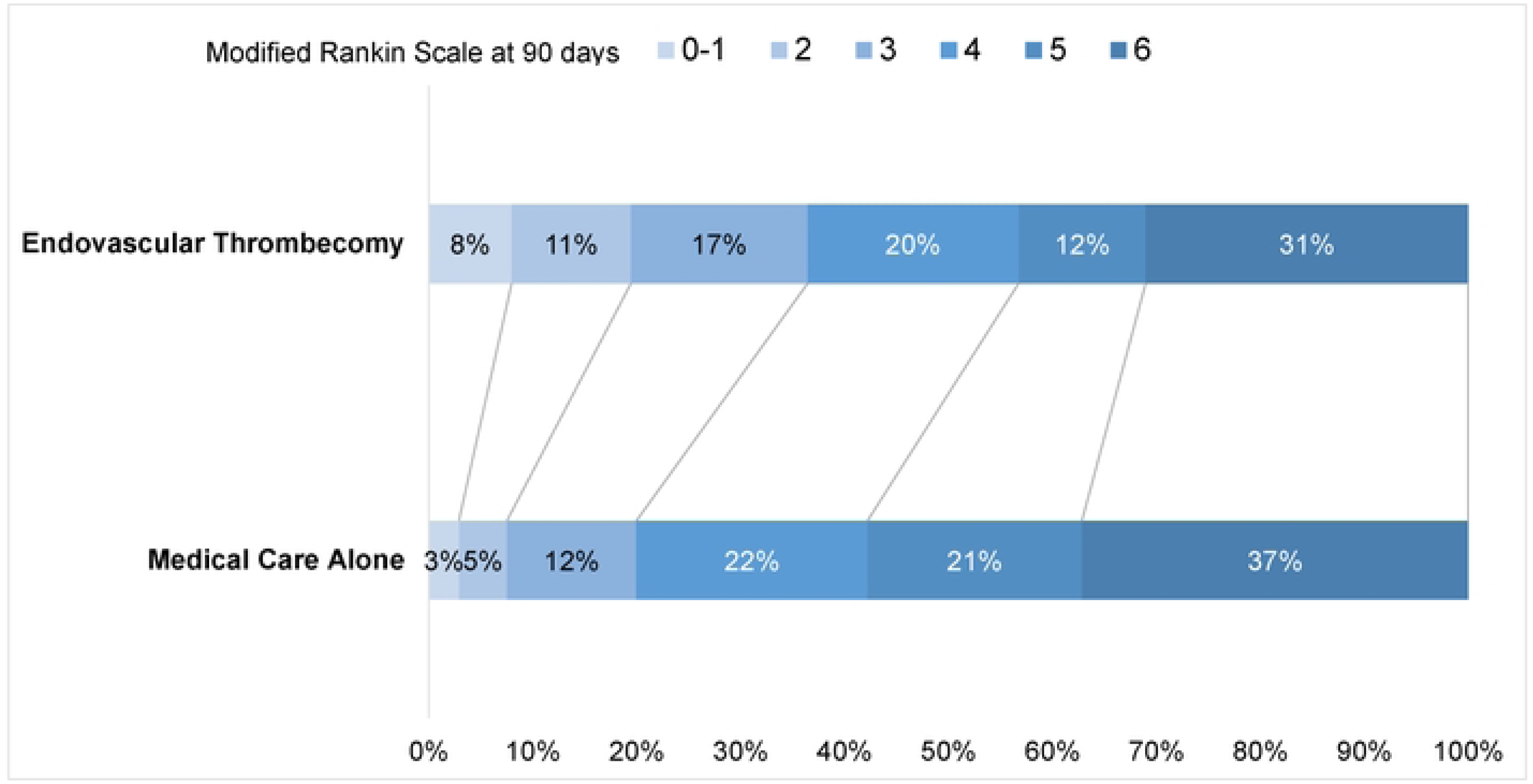
Distribution of Modified Rankin Scale Scores at 90 Days. A modified Rankin scale score of 0 to 1 indicates nondisabled, 2 disabled but independent, 3 dependent but ambulatory, 4 not ambulatory but not requiring constant care, 5 requiring constant care, and 6 death.

#### Ambulation (mRS score of 0 to 3) at 90 days

Pooled results showed that EVT compared with medical care alone was associated with a higher proportion of patients with ambulation at 90 days in AIS-LVO patients with large ischemic cores (6 trials; absolute risk, 36.5% vs 19.9%; RR 1.92, 95% CI 1.50 to 2.44, P < .001; NNT, 6).^5–10^ This represented 183 more ambulatory patients per 1000 treated with EVT compared with medical care alone. There was substantial heterogeneity among included trials (*P* for heterogeneity = .06; *I*^2^ = 54%) (eFigure 3 in Supplement).

**Figure 3.**
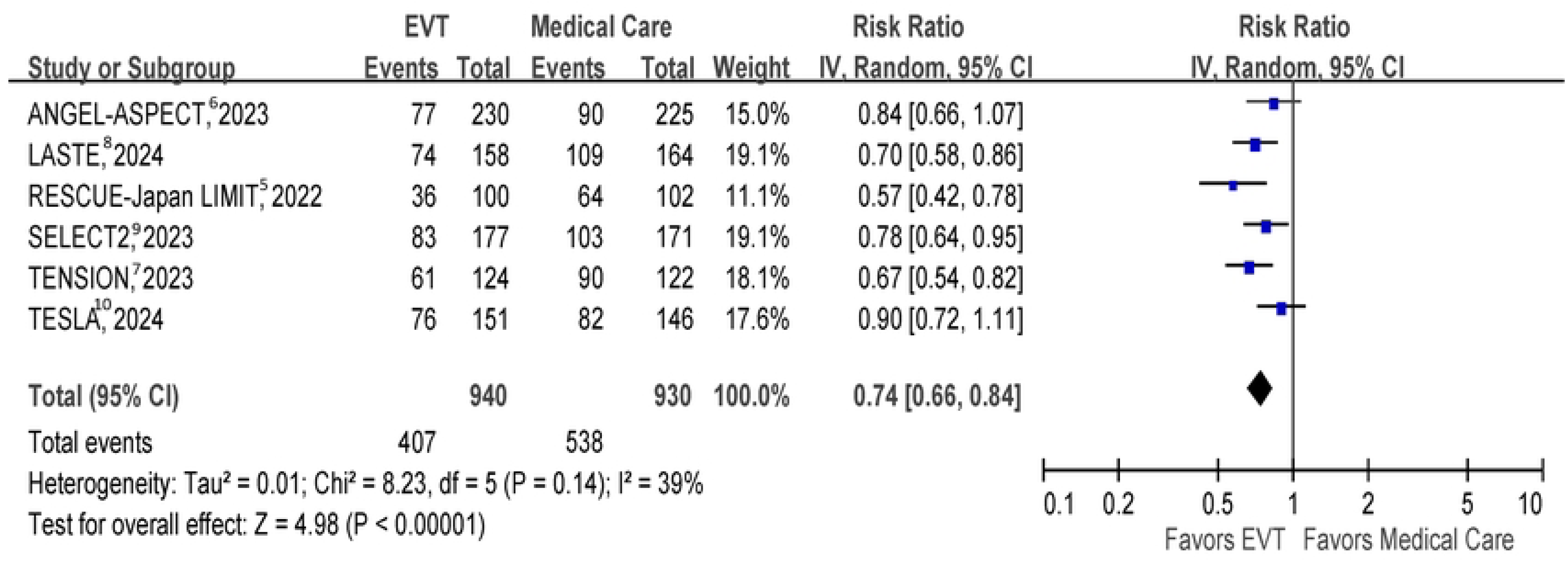
Requiring constant care or death at 90 days. Risk ratio with 95% confidence interval of requiring constant care or death, defined as modified Rankin Scale 5 to 6, with EVT plus medical care compared with medical care alone in AIS-LVO patients with large ischemic cores at 90 days. AIS-LVO, acute ischemic stroke due to large-vessel occlusions; EVT, endovascular thrombectomy.

#### Being nondisabled (mRS score of 0 to 1) at 90 days

Pooled results showed that EVT compared with medical care alone was associated with a higher proportion of nondisabled patients at 90 days in AIS-LVO patients with large ischemic cores (6 trials; absolute risk, 8.0% vs 2.9%; RR 2.64, 95% CI 1.71 to 4.08, P < .001; NNT, 21).^5–10^ This represented 47 more nondisabled patients per 1000 treated with EVT compared with medical care alone. Heterogeneity was not important among included trials (*P* for heterogeneity = .73; *I*^2^ = 0%) (eFigure 4 in Supplement).

#### Early neurologic improvement

The definition of early neurologic improvement varied among included trials. Early neurologic improvement was defined as a reduction of at least 8 points in the NIHSS score at 48 hours in the RESCUTE-JAPAN LIMIT,^5^ at 24 hours in the SELECT2,^9^ and 0-2 NIHSS score or reduction of at least 8 points at 6 days or discharge in the TESLA trial^10^ and defined as a reduction of at least 10 points in the NIHSS score or NIHSS score of 0 or 1 at 36 hours in the ANGEL-ASPECT^6^ and reduction of at least 10 points in the NIHSS score from the time of presentation to a thrombectomy-capable center to either day 7 or discharge in the LASTE trial.^8^

Pooled results of the five included trials that reported this outcome showed that EVT compared with medical care alone was associated with a higher proportion of patients with early neurologic improvement in AIS-LVO patients with large ischemic cores (5 trials; absolute risk, 18.6% vs 7.8%; RR 2.35, 95% CI 1.79 to 3.10, P < .001; NNT, 9).^5,6,8–10^ This represented 106 more patients with early neurologic improvement per 1000 treated with EVT compared with medical care alone. Heterogeneity was not important among included trials (*P* for heterogeneity = .41; *I*^2^ = 0%) (eFigure 5 in Supplement).

### Safety Outcomes

#### Requiring constant care or death at 90 days (mRS score of 5 to 6)

Pooled results showed that EVT compared with medical care alone was associated with a lower proportion of patients requiring constant care or dead at 90 days in AIS-LVO patients with large ischemic cores (6 trials; absolute risk, 43.3% vs 57.8%; RR 0.74, 95% CI 0.66 to 0.84, P < .001; NNT, 7).^5–10^ This represented 150 fewer patients requiring constant care or dead per 1000 treated with EVT compared with medical care alone. There was moderate heterogeneity among included trials (*P* for heterogeneity = .14; *I*^2^ = 39%) (Figure 3).

#### Death within 90 days

Pooled results showed that EVT compared with medical care alone was not associated with a significantly reduced risk of death within 90 days in AIS-LVO patients with large ischemic cores (6 trials; absolute risk, 31.5% vs 36.8%; RR 0.86, 95% CI 0.72 to 1.02, P =.08)^5–10^ and there was moderate heterogeneity among included trials (*P* for heterogeneity = .11; *I*^2^ = 45%) (eFigure 6 in Supplement).

#### Early symptomatic intracranial hemorrhage

Among included trials, the early symptomatic intracranial hemorrhage was defined either according to the Safe Implementation of Thrombolysis in Stroke Monitoring Study (SITS-MOST) criteria^19^ in 4 trials ^5,8–10^ or according to the Heidelberg bleeding classification^20^ in 3 trials.^6–8^ The LASTE trial reported early symptomatic intracranial hemorrhage using both the SITS-MOST criteria and the Heidelberg bleeding classification.^8^ For our pooled analysis, we chose to use the outcomes based on the SITS-MOST criteria.

Pooled results showed that EVT compared with medical care alone was not associated with a significantly higher proportion of patients with early symptomatic intracranial hemorrhage in AIS-LVO patients with large ischemic cores (6 trials; absolute risk, 4.4% vs 2.7%; RR 1.65, 95% CI 1.00 to 2.70, P =.05).^5–10^ Heterogeneity was not important among included trials (*P* for heterogeneity = .75; *I*^2^ = 0%) (eFigure 7 in Supplement).

#### Sensitivity Testing

Sensitivity test excluding individual trials serially yielded pooled results similar to the overall pooled estimates of the primary outcome, indicating that no single study had a disproportionate influence on the overall effect size.

#### Publication bias

The funnel plots showed no major asymmetry except for a small degree of publication bias, with a slight under-representation of a small study showing neutral effects (eFigure 8 in Supplement).

## DISCUSSION

Our systematic review and meta-analysis, which included data from six randomized clinical trials involving 1,870 individuals, demonstrated that for AIS-LVO patients with large ischemic cores in the anterior cerebral circulation within 24 hours of last known well time, compared to medical care alone, EVT plus medical care was associated with clinical benefit. Specifically, EVT plus medical care was linked to better functional outcomes, including increased rates of functional independence, reduced disability, improved ambulation, and early neurological improvement. For every 1000 patients, EVT led to 270 patients having less disabled outcomes, including 113 more independent patients than medical care. Additionally, this approach reduced the risk of requiring constant care or death at 90 days.

The magnitude of benefit of EVT for AIS-LVO patients with large ischemic cores in this study was less than that previously demonstrated for patients with small to moderate cores. For every 1000 patients, EVT led to 270 patients having less disabled outcomes in large core patients compared with 380 having a less disabled outcome in small to moderate core patients.^1^ The locus of benefit along the disability spectrum also differed, with benefit for nondisabled (mRS 0-1) and independent (mRS 0-2) outcomes greater for small to moderate core patients but benefit for ambulatory (mRS 0-3) and self-care capable (mRS 0-4) outcomes similar across core sizes. For example, for small to moderate vs large core patients, the increase in mRS 0-2 outcomes per 1000 patients was 195 vs 113, while for mRS 0-3 outcomes the increase was 200 vs 183.^1^ This diminished benefit in extensive ischemic cores, especially for the highest functional outcomes, likely stems from patients having already sustained a large burden of irreversible brain injury that cannot be salvaged despite successful reperfusion with EVT. Albeit with diminished benefits compared to smaller cores, EVT may be still justified for use in AIS-LVO patients with large ischemic cores over medical care alone because of potentially improving functional outcomes and decreasing the risk of very poor outcomes at 90 days.

In EVT-treated patients in an individual participant level analysis of a subset of the completed trials, clinical outcomes worsened as ischemic injury estimates increased.^21^ Our study-level meta-analysis could not define the lower boundary of ASPECTS value for a clinical benefit of EVT. A meta-analysis of individual patient-level data did not find any imaging biomarker that could be used as a treatment effect modifier to decide which AIS-LVO patients should be treated with EVT or not.^22^ Furthermore, since preservation of the high cortical regions was more strongly associated with improved outcomes compared to the deep regions,^23^ not all low ASPECTS infarcts are highly disabling infarcts. Also, initial CT perfusion may overestimate final infarct core, and selecting patients for EVT based on the CT perfusion mismatch concept may deny treatment to patients who might still benefit from reperfusion.^24^ About 15% to 25% of patients had permanent reversal of DWI after EVT, associated with good clinical outcome, which highlight the pitfalls of DWI to define ischemic core.^25,26^ Also, an observational study showed that recanalization was associated with improved functional outcome in patients without salvage tissue on CT perfusion.^27^ A recent meta-analysis of a patient with ASPECT score value of 0 to 2 in the ANGEL-ASPECT trial^6^ and the SELECT2^9^ suggest that EVT might still be beneficial for patients with ASPECT score < 3^28^ and similar finding was observed in the post-hoc analysis of the TENSION trial.^7^ Therefore excluding AIS-LVO patients from EVT solely based on very low ASPECTS value or large ischemic core on CT perfusion or DWI may not be appropriate. Still, the RESCUE-Japan LIMIT suggested that EVT was cost-effective for participants with an ASPECTS 4 to 5, but not for those with an ASPECTS value of 3 or less from both the US and Japan perspectives.^29,30^ To determine the eligibility for EVT in AIS-LVO patients with large ischemic cores, it may be necessary to consider not only the quantity of infarct but also the location of the infarct and the quality and functioning of the non-infarcted brain tissue.

The primary outcome varied among the included trials and the TESLA trial^10^ was the only trial that failed to show superiority of EVT to medical care in its original primary outcome. Also, the TESLA trial did not show a significant benefit of EVT over medical care alone for improving functional independence, reducing disability, and reducing patients requiring constant care or dead.^10^ Successful reperfusion was only 73% in the TESLA trial^10^ whereas 80% or more of successful reperfusion was found in other trials.^5–8^ Since poor reperfusion after EVT was associated with worse outcomes compared with medical care alone,^31^ a relatively lower rate of successful reperfusion in the TESLA trial^10^ was likely to hamper the functional outcome in patients allocated to the EVT group.

### Limitations

This study has limitations. First, since this is a study-level meta-analysis, we were unable to conduct subgroup analyses because each trial reported its subgroup analysis based on its original primary outcome. An individual participant pooled analysis of all trials is warranted to solve this problem. Second, the level of ASPECTS value below which EVT should not be recommended is not known based on the evidence currently available. Comprehensive cost-effective analyses may be needed before EVT can be applied universally for all patients with AIS-LVO within 24 hours of last known well time. Third, the imaging modality to define large ischemic cores varied among included trials. The trial that predominantly relied on non-contrast CT scans for identifying large ischemic cores had enrolled patients within 12 hours of stroke symptom onset.^7^ Consequently, for selecting appropriate AIS-LVO patients

with large ischemic cores for EVT in cases where the last known well time falls between 12 and 24 hours, more advanced imaging techniques such as CT perfusion or MRI might be required. Fourth, per the original definition of large core, greater than one-third of the middle cerebral artery territory or 100 ml, the current analysis actually addresses combined larger moderate core and true large core patients, rather than large core patients alone.

### Conclusions

Our meta-analysis of randomized clinical trial data suggests that for patients with AIS-LVO and large ischemic cores, EVT combined with medical care may be associated with improved functional outcomes and reduced severe disability or death at 90 days compared to medical care alone. Further individual patient-level pooled analysis and cost-effectiveness analysis are desirable to determine the lower limit of the ASPECTS or the upper limit of infarct volume on CT perfusion or DWI that would make a patient most suitable for EVT.

## Data Availability

This is a meta-analysis of published randomized controlled trials, All data can be found at journal website of each included trial.

## ACKNOWLEDGMENT

**FUNDING/SUPPORT** This work was supported by Ministry of Science and Technology, Taiwan, grant number: MOST 108-2314-B-182-017-, MOST 109-2314- B-182-033-, MOST 110-2314-B-182-036 -MY2 and Chang Gung Memorial Hospital, Taiwan, grant number: CMRPG6H0191, CMRPG6H0441.

**ROLE OF FUNDER/SPONSOR STATEMENT** The sponsors played no role in the design and conduct of the study; collection, management, analysis, and interpretation of the data; preparation, review, or approval of the manuscript; and decision to submit the manuscript for publication.

**ACCESS TO DATA AND DATA ANALYSIS** Meng Lee had full access to all the data in the study and takes responsibility for the integrity of the data and the accuracy of the data analysis.

Chun-Hsien Lin, Meng Lee, and Borja Sanz-Cuesta are responsible for the data analysis.

**Conflict of Interest**

The authors have no competing interest.

## Notes

### Competing Interest Statement

The authors have declared no competing interest.

### Funding Statement

Yes

### Author Declarations

This is a meta-analysis of published RCTs and exempt from IRB.

